# Missorting of Plasma miRNAs in Aging and Alzheimer’s Disease

**DOI:** 10.1101/2022.03.18.22272622

**Authors:** Maria Čarna, Jan S. Novotny, Neda Dragišič, Hanuš Slavik, Kateřina Sheardova, Yonas E. Geda, Martin Vyhnalek, Jan Laczo, Jakub Hort, Zixu Mao, Robert A. Rissman, Marian Hajduch, Eric B. Dammer, Gorazd B. Stokin

## Abstract

Aging is the greatest risk factor for Alzheimer’s disease (AD)^1^, a pervasive cognitive disorder with unsettled etiology. The precise role of aging in AD, however, remains poorly understood. Accumulating evidence documents the dysregulation of circulating microRNAs (miRNA) separately in aging^2,3^ and AD^4^. Considering miRNAs play a role in aging and longevity^5,6^, we comprehensively test which aging-associated miRNA changes are observed in AD, and change in AD beyond aging in the circulating miRNA network. Here we show that plasma miRNAs in aging are downregulated and preferentially targeted to extracellular vesicle (EV) content, while in AD, miRNAs are further downregulated and of exclusive EV origin. We further show that miRNAs in AD display altered proportions of motifs relevant to their loading into EVs^7,8^ and secretion propensity^9^. Considering endosomes play a role in the genesis of EVs^10,11^ and are compromised early in AD^12^, these findings implicate endosomal pathology underlying the AD plasma miRNA profile and its potential as a mechanistic AD biomarker. Striking similarities between plasma miRNA profiles in aging and AD further suggest that the AD miRNA network profile reflects a pathological exacerbation of the aging process whereby physiological suppression of AD pathology by plasma miRNAs becomes insufficient.

## Introduction

MicroRNAs (miRNAs) are non-coding RNA molecules (ncRNA) involved in post-transcriptional gene silencing^13^. In humans, RNA interference regulates approximately 60% of gene transcripts^14^. Since 2007, many studies have reported miRNAs in various body fluids including blood^15^. These circulating miRNAs are released from cells either free or packaged into exosomes and other EVs or bound to carriers such as Argonaute 2 (AGO2)^16^. Packaging of miRNAs into exosomes takes place in the endosomal compartment^10^ and involves interplay between miRNA motifs dictating their secretion, such as the EXOmotifs^9^, as well as sorting which includes recognition motifs for heterogeneous nuclear ribonucleoproteins (hnRNPs)^7^ and AGO2^8^. Cells secrete miRNAs to maintain their homeostasis and intercellular communication^17^, but also release miRNAs in response to injury^18^. Therefore, while playing a role in regulating gene expression, miRNAs have the potential to serve as minimally invasive biomarkers and offer insight into novel therapeutic approaches for conditions like AD.

Prior studies reported dysregulated miRNAs in the blood in AD with the majority of miRNAs underrepresented and frequently targeting amyloid precursor protein, its fragments and proteolytic machinery, all intimately linked to the pathogenesis of AD^4,19^. Intriguingly, blood cell miRNAs in aging were also found to be underrepresented^2,3^. These observations are suggestive of a relationship between miRNAs, aging and AD, which demands further investigation. To investigate the cross-talk between aging and AD at the miRNA level, we compared plasma miRNA expression patterns of healthy young and old individuals and patients with AD. Our study leverages co-expression profiling of plasma miRNAs, permitting insights into AD mechanisms, and definition of aging and AD associated miRNA profiles, opening the possibility to determine specific miRNA roles in AD.

## Main text

### Surveying plasma ncRNAs in aging and AD

To gain insight into ncRNAs in aging and AD, we examined plasma harvested from young and old individuals and patients with AD (Tbl. S1A). Plasma ncRNAs consisted of miRNAs, RNA stem-loops, small nucleolar RNAs (snoRNAs), box C/D RNAs and box H/ACA RNAs in addition to traces of small Cajal-body specific RNAs (scaRNAs) and 5.8S rRNAs (Fig. 1A, Tbl. S1B). Principal component analysis (PCA) and heatmap found that miRNAs of old individuals and patients with AD cluster close to each other and segregate significantly from young individuals (Fig. 1B, C, Fig. S1A-C, Tbl. S1C). Old individuals and patients with AD also demonstrated predominant downregulation of differentially expressed (DE) miRNAs compared with young individuals (Fig. 1D, Tbl. S1D). This was particularly evident in patients with AD where no DE miRNA showed upregulation compared with old individuals.

**Figure 1.**
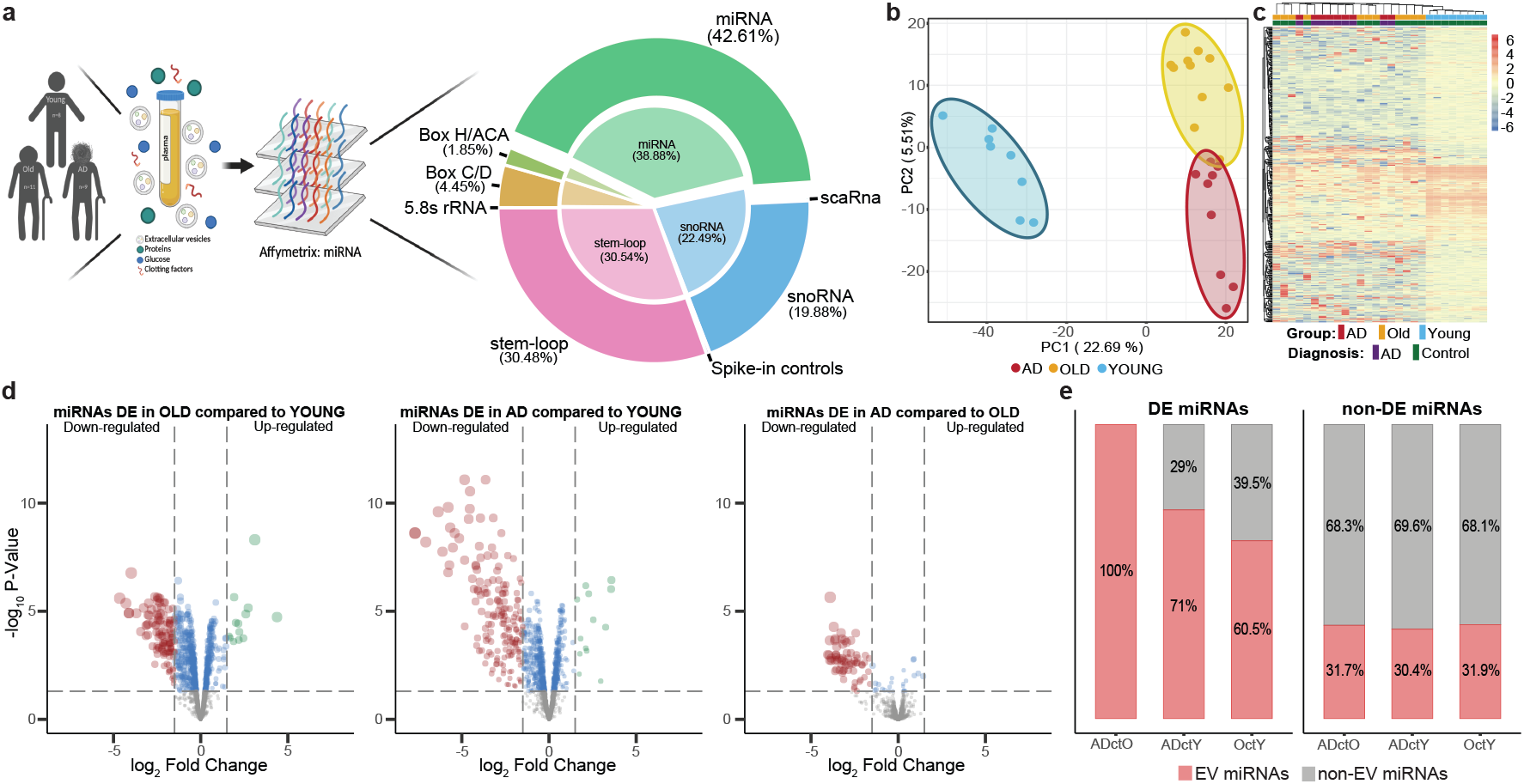
Plasma ncRNAs in aging and AD. **(A)** Experimental design and composition of plasma ncRNA. Inner pie displays absolute number of individual types of probes, outer donut shows number of unique probes, **(B)** PCA of miRNAs in individual experimental groups, **(C)** Heatmap of hierarchically clustered top 500/2578 miRNAs with the greatest in-row variability and annotation for individual experimental groups, **(D)** Volcano plot displaying the log_2_ fold change (*x* axis) against the −log_10_ statistical *P* value (*y* axis) for DE miRNAs between individual experimental groups (*P* value cut-off drawn at equivalent of FDR adjusted *P*=0.05, fold-change cut-off drawn at ±1.5), **(E)** Vesiclepedia-based proportions of miRNAs found in EVs in individual experimental groups (chi-square test for DE groups and DE versus non-DE groups, p-value < 0.01).

Considering Vesiclepedia allows searching for EV cargoes including miRNAs^20^, we next investigated proportions of miRNAs included in EVs (Fig. 1E). Approximately 30% of non-DE miRNAs were present in EVs across all the comparisons between the experimental groups. This contrasted with the significantly higher proportions of DE miRNAs present in EVs, especially in patients with AD compared with old individuals. Therefore, in aging, downregulated miRNAs are more common in EVs, while in AD, these age-related changes become exacerbated.

### Aging and miRNA modules in AD

To investigate unbiased clusters of co-expressed miRNAs and their correlation to aging and AD, we performed weighted gene co–expression network analysis (WGCNA). Hierarchical clustering with dynamic tree cutting of the scale-free miRNA co-expression network identified miRNA modules associated with sample traits including age, sex, cognition (MMSE) and AD status (Fig. 2A, Tbl. S2B). Turquoise followed by blue module eigenvalues correlated most significantly with age, while purple followed by pink and blue module eigenvalues demonstrated best correlation with AD status (Fig. 2B). Testing for robustness of the miRNA network modules by comparison with an independent dataset^21^ found good preservation of the blue (Zsummary>2) and purple (Zsummary=10) modules (Fig. S2A, B). Therefore, we identified two stable modules related to the effects of aging and AD. These two modules showed the most significant enrichment in overrepresented biological processes in the pathway analysis based on validated miRNA targets (Fig. 2C, Tbl. S2C). While only the blue module exhibited significant enrichment for GWAS-derived AD risk factors (Fig. 2D, Tbl. S2D), the purple module showed highest percentage of miRNA targets related to AD and involving APP (Fig. 2E, Tbl. S2E, Fig. S3, Extended Data Tbl. C).

**Figure 2.**
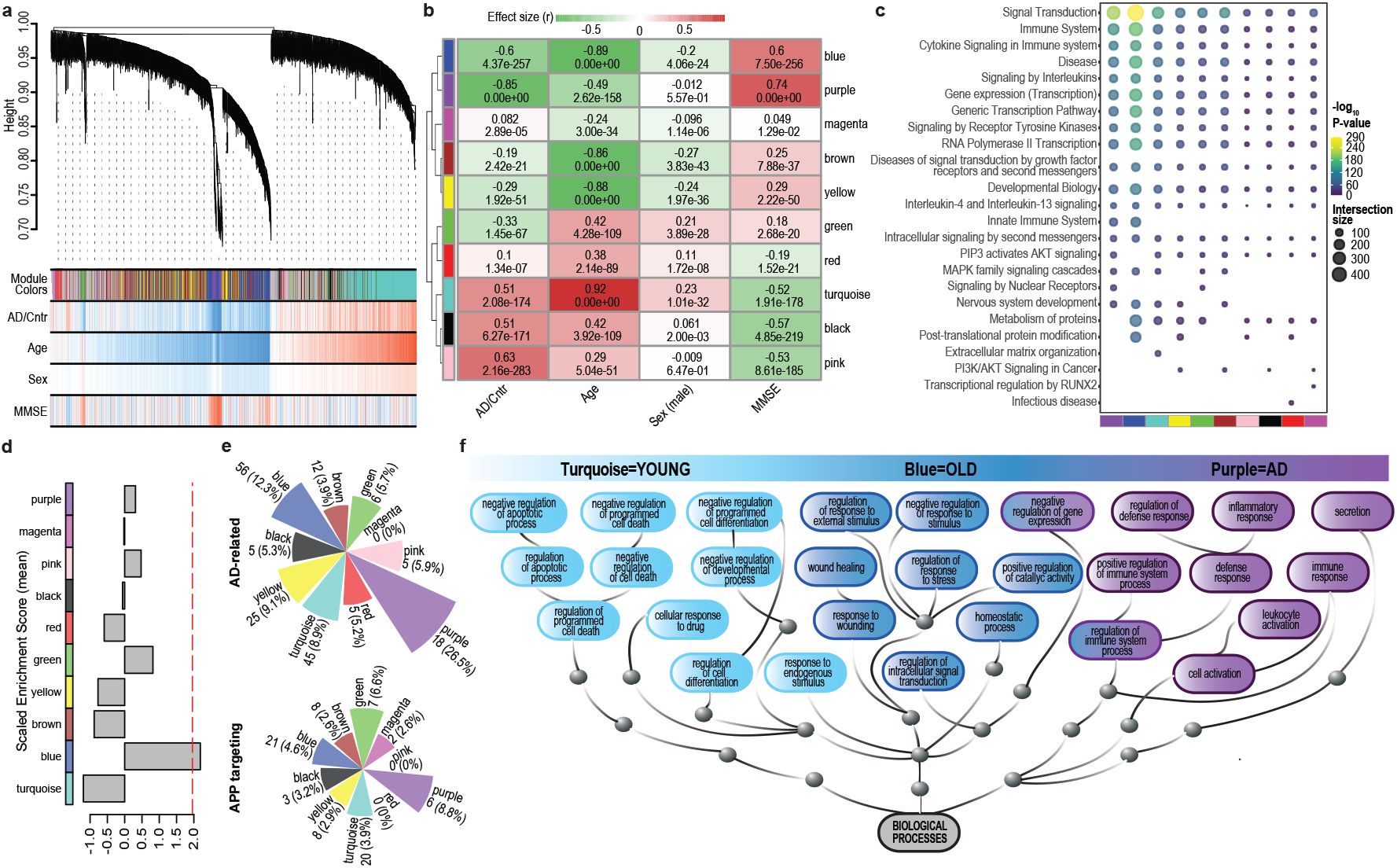
Characteristics of plasma miRNA WGCNA network in aging and AD. **(A)** WGCNA module distribution in a hierarchically clustered dendrogram based on co-expression and correlation of miRNAs and sample traits, **(B)** Heatmap of correlations between WGCNA module eigenvalues of co-expressed miRNAs and sample traits, (**C**) The most significant REACTOME-pathways of mature miRNA targets assigned to individual modules, (**D**) Enrichment for GWAS-derived AD risk factors in different modules. The dashed red line indicates Z score of 1.96 (*P*=0.05), (**E**) Pie-charts of the count and percentages of miRNAs in individual modules related to AD (upper) and targeting APP (bottom), (**F**) An acyclic graph showing significantly enriched GO categories of biological processes depicting top 10 GO terms of HUB miRNA targets for turquoise (young), blue (old), and purple (AD) module.

We last delineated biological processes that represent unique changes in AD over human lifetime based on the targets of miRNAs assigned to age- and AD-correlated modules (Fig 2F, Tbl. S2F). In young, turquoise module miRNAs control targets negatively regulating apoptosis and cell death, while with aging these processes become more silenced. The blue module miRNAs in young repress gene products involved in stress and wound healing and repression become less controlled with aging. In contrast, purple modules miRNAs fail to regulate inflammation and immune response in AD.

### Characteristics of miRNA modules in AD

To test for consistency, we compared DE plasma miRNAs identified in AD with those previously reported in AD, Parkinson’s disease (PD), amyotrophic lateral sclerosis (ALS) and fronto-temporal dementia (FTD) (Fig. 3A, references in materials and methods). Comparison showed preponderant downregulation of plasma miRNAs across all disorders and identified 11 plasma miRNAs DE exclusively in AD in this and previous studies (Fig. S4, Tbl. S3A). Most miRNAs belonged to the purple module including hsa–miR-106a-5p and hsa–miR-423-3p, which were not reported previously in studies selected for this comparison (Fig. 3B).

**Figure 3.**
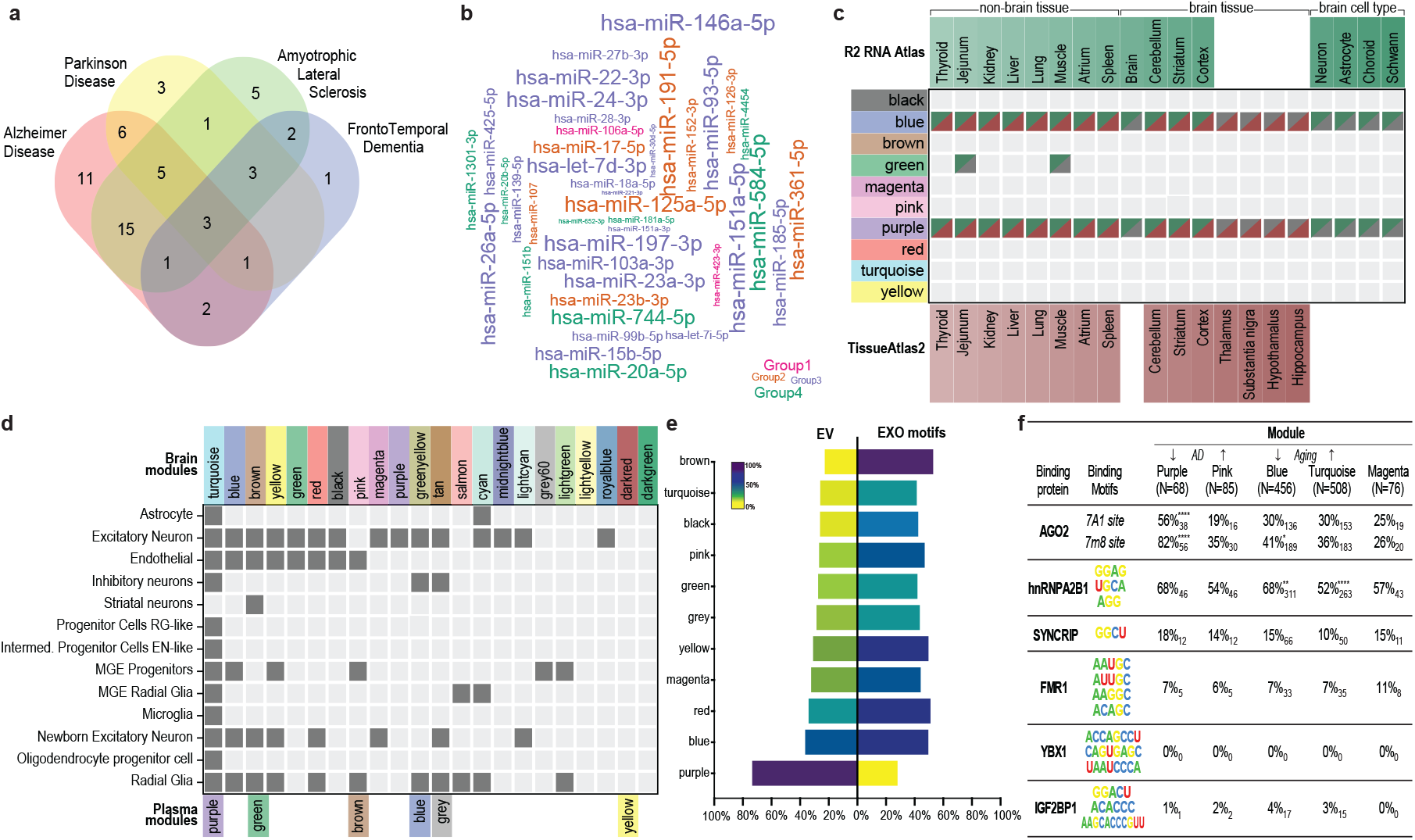
Specificity, origin and features of plasma miRNA modules in AD. (**A**) Venn diagram showing DE miRNAs in AD and other neurodegenerative disorders in our and previous studies (FDR adjusted *P*=0.05, fold-change ±1.5), (**B**) Word cloud of DE purple module miRNAs in AD in our and previous studies. Individual groups include DE miRNAs in AD **1)** in our study only, **2)** in our and previous studies, **3)** in our study and in AD and other disorders in previous studies, **4)** in our study and in other disorders in previous studies (FDR adjusted *P*=0.05, fold-change ±1.5, asterisks indicate unreported miRNAs in the brain based on TissueAtlas), (**C**) Overlap between plasma modules and miRNAs in tissues and cell-types in R2 RNA and the Tissue Atlases. Colour scale shows the number of miRNAs overlapping in each tissue and cell-type. Only miRNA meeting minimal abundance criterion of 10 reads were included, (**D**) Heatmap showing plasma modules significantly overlapping with brain module miRNAs (-log^10^ *P* value>1.301), (**E)** Vesiclepedia-based proportions of miRNAs found in EVs and harbouring EXO motifs in individual modules, **(F)** Count and proportion of miRNAs in the purple and select other modules containing sequence motifs for RNA-binding proteins, **(F)** Proportion of miRNAs containing sequence motifs for RNA-binding proteins in different modules (counts shown as subscript). Stars indicate significance level (*<0.05, **<0.01, ****<0.0001).

Since AD is characterized by brain pathology, we asked whether DE plasma miRNAs in AD exist also in the brain. Comparison of miRNA modules with miRNAs residing in tissues catalogued in the R2 RNA^22^ and the Tissue Atlases^23^ showed that purple module miRNAs were enriched in the brain, but expressed also in other tissues, while the blue module miRNAs showed no differences in tissue expression (Fig. 3C, Tbl. S3B). Approximately 45% of the plasma miRNAs could not be matched to any tissue. We also compared plasma with previously described brain miRNA modules^24^ and identified several matches with the purple module showing extensive presence across brain cell populations (Fig. 3D, Tbl. S3C).

We next investigated whether miRNA modules correlating with AD demonstrate increased proportion of miRNAs in EVs. Unlike all the other modules, the majority of purple module miRNAs were reported in EVs (Fig. 3E, Tbl. S3D). Considering secretion^9^, but also packaging of miRNAs^7,8^ is regulated by sequence motifs, we last examined proportions of sequence motifs in different modules. Proportion of EXOmotifs was significantly reduced in the purple module, AGO2 binding motifs were significantly enriched in the purple and to a lesser extent in the blue module, while hnRNP A2B1 motifs were significantly enriched in the blue and turquoise age-related modules (Fig. 3E, F; Tbl. S3E, S3F). Altogether, miRNAs specific to AD exhibit a disproportionate amount of sequence motifs, are overrepresented in EVs and reside in the brain as well as in other tissues.

### Modeling aging with AD

To probe the effects of aging on miRNAs in AD further, we compared DE miRNAs and their targets across all the comparisons between the experimental groups (Fig. 4A). There were no DE miRNAs unique to AD compared with either young or old individuals. A total of 908 miRNAs were DE in AD and also in old compared with young individuals (Tab. S4A). These miRNAs reflected healthy aging with their targets involved primarily in synaptic transmission. Only 89 miRNAs were DE in old versus young and in AD versus young or old. These miRNA changes were consistent with aging and specific for AD with their targets involved in chromatin regulation, regulation of the cell cycle and RNA and proteasomal catabolism (Fig. 4B; Tbl. S4A, S4B).

**Figure 4.**
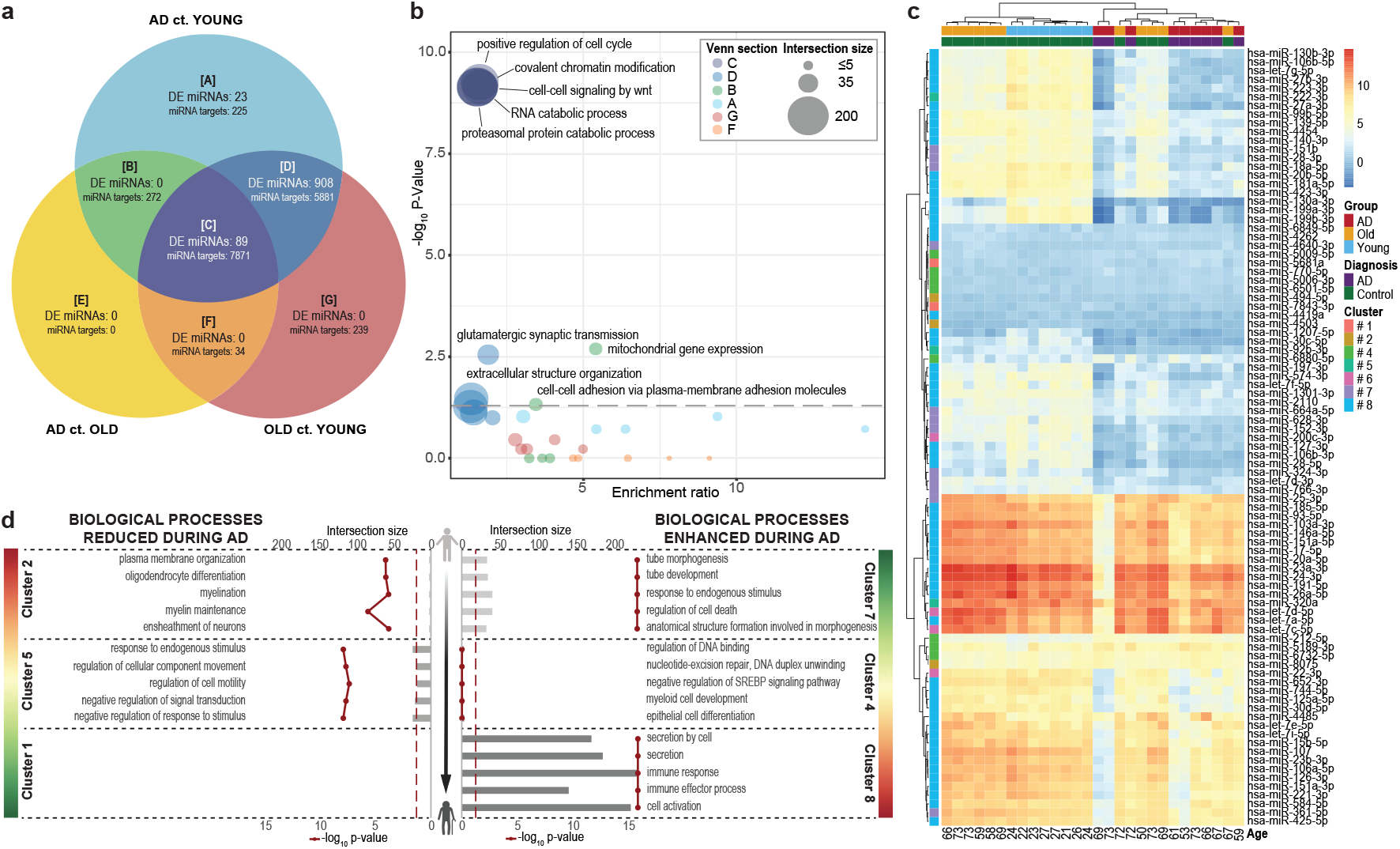
The effects of aging on plasma miRNAs in AD. (**A**) Venn diagram showing DE miRNAs and their targets between experimental groups (FDR adjusted P=0.05), (**B**) GO terms of overlapping miRNA targets in the Venn diagram. The colour and size of the bubbles represent Venn diagram section and intersection sizes, respectively, (**C**) Hierarchically clustered heatmap showing expression of 89 DE miRNAs in all experimental groups. Vertical annotation shows partitioning of miRNAs into subclusters based on the relative positions of their DE vectors in aging and AD (subclusters 1-8), (**D**) GO enrichment analysis of biological processes for the targets of dysregulated miRNA subclusters during the aging. Upper X-axis and grey horizontal bars show the number of targets involved in biological processes, bottom X-axis and vertically connected red dots show -log^10^ P value of individual GO terms. Lateral colour scales show involvement of biological processes in aging (green: less involved, red: more involved).

Age-dependent and AD specific miRNAs segregated into 8 subclusters (Fig. 4C, Fig. S5, Tbl. S4C). To find out the functional outcomes of aging and AD, we performed enrichment analysis to identify key changes in biological processes based on the interaction between aging and AD miRNAs and their targets in individual subclusters. Modeling revealed that aging and AD perturb several processes previously described to play a role in AD including oligodendrocyte homeostasis^25^, subcellular component movement^26^, DNA repair^27^ and immune response^28^ (Fig. 4D, Tbl. S4D).

## Conclusions

Aging is the greatest risk factor underlying AD, however, the mechanisms by which aging plays a role in AD remain enigmatic. We establish a relationship between plasma miRNAs, aging and AD by showing progressive reduction of plasma miRNAs in aging and furthermore in AD. This relationship is consistent with decreased blood miRNA levels reported separately in aging^2,3^ and AD^29,30^ and suggests that in terms of plasma miRNAs, AD represents a pathological exacerbation of physiological aging. Observed impairment in secretion and/or degradation of circulating miRNAs suggests that physiological mechanisms of post-transcriptional gene silencing become insufficient in AD. Progressive impairment of translational repression regulating protein expression during aging therefore contributes to the pathogenesis of AD also by perturbing APP levels^19,31^ and producing downstream effects akin to increased *APP* gene dosage described in a subset of AD cases^32,33^.

The most striking result, however, is that miRNAs which decrease in AD plasma in a coordinated, co-expressed pattern with high significance of correlation to AD status as a trait are found exclusively in EVs with perturbed proportions of sequence motifs suggesting that circulating miRNA packaging, secretion and/or uptake are compromised in AD. Considering endosomes participate in exosome formation^10,11^ and are invariably affected early in AD^12^, it is plausible to hypothesize that plasma miRNA changes in AD are the result of endosomal pathology. This hypothesis is further corroborated by genetic and biochemical evidence of endosomal alterations in aging and AD^34-36^. Besides these AD-related miRNAs (purple module) reflecting endosomal pathology, we also identified aging and AD correlating miRNAs (blue module) that associate with GWAS-derived risk factors of AD and mirror events implicated in the pathogenesis of AD. Further experiments and data are needed to elucidate the origin, targets and signalling pathways of miRNAs involved in AD. This will also clarify whether all plasma miRNA changes in AD reflect brain pathology and/or alternatively, whether the brain in AD serves as a substrate for miRNA signals from other tissues.

## Supporting information

Tbl. S1

Tbl. S2

Tbl. S3

Tbl. S4

Extended Data Tbl.

## Data Availability

All data produced in the present work are contained in the manuscript

## Materials and Methods

### Samples and clinical data

Plasma samples and clinical data were obtained through the Shiley-Marcos Alzheimer’s Disease Research Center at the University of California San Diego in addition to the commercial vendor, PrecisionMed, and the Czech Brain Aging Study^1^ funded by the International Clinical Research Center of St. Anne’s University Hospital and running in the Departments of Neurology at Motol and St. Anne’s in Prague and Brno, respectively. Plasma samples analysed and presented in this study were collected from 24,3±2,2 years old healthy young subjects (n=8), 66,3±7,4 years old healthy old subjects (n=11) and age-matched 65,9±6,8 years old patients with AD (n=9, Tbl. S1A). In brief, AD was diagnosed following the National Institute of Neurological and Communicative Disorders and Stroke (NINCDS) – Alzheimer’s Disease and Related Disorders Association (ADRDA) clinical criteria^2,3^. Clinical data of the diagnostic work-up encompassed behavioral and cognitive testing including Mini-Mental State Examination (MMSE), general and neurological clinical assessments, blood exams, brain imaging to exclude mass lesions and cerebrospinal fluid amyloid 1-42 peptide and total tau measurements.

Plasma samples and clinical data were collected following Institutional Review Board approvals with written consent obtained from all subjects. Competency forms for all patients with AD were signed by physicians following evaluation of their capacity to sufficiently understand tests and examinations involved in the research and their capability of consenting on their own behalf.

### Plasma RNA Isolation

RNA was isolated from plasma samples using the miRNeasy kit (Qiagen, Germany) according to the manufacturer’s instructions with the following modifications. Plasma samples were mixed with five sample volumes of QIAzol reagent, vortexed for 10 s and then incubated at room temperature for 5 min to inactivate RNases. The next steps were performed according to the manufacturer’s protocol. RNA extracted from plasma was concentrated to a minimum volume of approximately 10 μl and quantified using Nanodrop 1000 (Thermo Fisher Scientific, USA).

### MiRNA Microarray

Extracted plasma RNA was used for poly A tailing and ligation with the FlashTag Biotin HSR RNA Labelling Kit (Affymetrix, Santa Clara, CA, USA). The labelled samples were mixed with a hybridization cocktail (2×Hybridization Mix, 27.5% formamide, DMSO, 20× Eukaryotic Hybridization Controls, Control Oligonucleotide B2 from GeneChip Eukaryotic Hyb Control Kit, Affymetrix) and hybridized onto a miRNA 4.0 Array (Affymetrix, USA) at 48 ^°^C for 16 h. Thereafter, washing and staining were performed using an Affymetrix GeneChip Fluidics Station 450. Scans were performed using an Affymetrix GeneChip Scanner 3000. Data analysis was performed using the Affymetrix miRNA Array QC tool. Experiment was performed following the manufacturer’s instructions.

### Data processing

Raw signals from microarray GeneChip (.CEL files) were normalized with RMA-DABG spike-in human-only normalization using the Transcriptome Analysis Console (TAC v4.0). Subsequently, duplicate probes were removed and only probes with maximum variance being retained using the collapseRows function of the WGCNA package in R^4^. Only mature miRNAs were filtered to form the final dataset.

### Differential expression analysis

Pairwise differentially expressed proteins were identified using the Student’s t test followed by the Benjamini-Hochberg (BH) FDR correction. The analysis was performed using the limma package in R^5^. The results were plotted using volcano plots generated by the ggplot2 package. A threshold of adj. p-value < 0.05 (or -log^10^ P-value>1.301) and log^2^ fold change ±1.5 was used to identify differentially expressed miRNAs, except for the analysis of biological consequences of aging on plasma miRNAs in AD, where only a threshold of adj. p-value < 0.05 was used (Tbl. S1D, Extended Data Tbl. A).

### Weighted gene co-expression analysis

Weighted Gene Co-expression Network Analysis (WGCNA) was used to detect groups of co-expressed miRNAs using WGCNA::blockwiseModules function^6^. The following WGCNA network configuration was adopted: network type=signed, soft threshold power=8, deepSplit=4, merge cut height=0.07, minimum module size=10, mean TOM denominator, correlation type=weighted bi-correlation (bicor) and reassign threshold=0.05 with partitioning about medoids respecting the dendrogram stage included. The assignment of miRNAs to modules was further refined by iterative re-assignment until all module members had a kMEintramodule value ≥ 0.28, with kMEs also calculated via bicor (Tab. S2A). The obtained network and module correlations with participants’ clinical traits (age, AD status, sex, MMSE score) were plotted using the WGNCA::plotDendroAndColors function and pheatmap packages.

### Modules preservation

To verify the stability of the individual modules, we computed module preservation using the WGCNA::modulePreservation function (with 200 permutations)^7^. Using Z-summary and MedianRank statistics, we quantified how density and connectivity patterns of modules defined in a reference data set (current study) are preserved in a comparative data set^8^ (Extended Data Tbl. B). The comparative data set included 10 microarray-derived plasma samples (5 AD and 5 age-matched healthy samples) and 1945 miRNAs of which 1793 overlapped with the miRNAs in the current study.

### miRNA target prediction

miRTargetLink 2.0 (https://ccb-compute.cs.uni-saarland.de/mirtargetlink2/) working with miRTarBase 8.0 from Homo sapiens was used to provide strongly validated miRNA targets^9^. The final set of corresponding target genes of miRNAs was established after removal of duplicate results.

### Gene Ontology (GO)

To examine the biological functions of the miRNA targets, we performed functional enrichment analysis using GO and REACTOME pathways databases. Functional enrichment of differentially expressed miRNAs and within the WGCNA modules was performed using WebGestalt (http://www.webgestalt.org) web tool^10^. The set of all targets identified in the hub miRNAs from correlated miRNA modules was used for over-representation analysis. Benjamini-Hochberg FDR correction was used to assess the significance level and a minimum of 5 genes per ontology were used as filters prior to pruning the ontologies. Hub miRNAs were selected based on the top greatest Module Membership (MM) values (kME >0.8). An integrated platform linking miRNAs, their targets and their functions, miRNet (https://www.mirnet.ca/) was used to predict the minimum downstream target genes of the screened AD related-miRNAs.

### GWAS module association

To determine if any protein products of GWAS targets were enriched in a particular module, we used previously published SNP summary statistics^11^ to calculate the gene-level association value using MAGMA (version 1.08b)^12^ as previously described^13^. A list of 1,822 genes with gene-based GWAS P < 0.05, including APOE, was used for enrichment analysis^14^.

### miRNA tissue and disease specificity analysis

Data from the R2Atlas (https://r2.amc.nl/) and the TissueAtlas v2^15^ were used to analyse the tissue and cell type specificity of miRNAs in individual WGCNA modules. The miRNA expression data for brain and non-brain tissue and cell-types were generated by NGS (with RPMM normalization) in both atlases. miRNA was considered to be present in respective tissue if it passed a minimal abundance criterion of 10 reads. Overlap analyses of module miRNAs with tissues were conducted by Fisher’s exact test and Jaccard index using GeneOverlap::newGOM package and function. To test for specificity of miRNAs and miRNA WGCNA module changes in AD, we compared DE miRNAs in our study with previous studies of DE miRNAs in AD^16,17,18,19,20,21,22,23^, PD^22,23,24,25^, ALS^23,26,27,28^ and FTD^23,29,30^ (Extended Data Tbl. D).

### Inclusion in EVs and sequence codes of plasma miRNAs

To test what proportions of miRNAs originate from EVs we calculated percentages of plasma miRNAs overlapping with the list of miRNAs previously identified in EVs^31^. To establish proportions of miRNAs harbouring EXO motifs, we calculated how many miRNAs carry EXO motifs in different WGCNA modules and experimental groups^32^. MiRNAs were considered harbouring an EXO motifs only if miRNA sequence contained exclusively EXO motifs or the number of EXO motifs in the sequence was greater than the number of cellular retention CELL motifs.

For five selected modules we also analysed whether their miRNA sequences contain sequence motifs required for their recognition and binding to select AD- and miRNA sorting-associated RNA-binding proteins^33,34^. Binding motifs for all RNA-binding proteins except AGO2 were extracted from their respective sources as miRNA-side sequences. For AGO2, AGO2-side sequence heptamers were extracted from the source. The reverse-complement sequence of the motif was used for analysis (Figure 3F and Tab. S3G). Enrichment of individual binding proteins in selected modules was examined using Fisher’s exact test.

### Statistics and visualization

All statistical analyses were performed in the RStudio environment (v.1.4.1717 with R v.4.1.1). The base prcomp function in R was used for Principal Component Analysis of the normalized expression data. Heatmaps of miRNA expression and overlap of miRNAs in modules to other modules and to tissue-specific lists were plotted using the pheatmap package. Bubble plots of biological functions and pathways, Venn diagrams and wordcloud were generated using the ggplot2, Venn and wordcloud packages, respectively.

## Acknowledgements

We thank past and current members of the Stokin Lab for support and feedback. We thank Drs Nowakowski and Kosik for providing script for cluster analysis.

## Author’s contributions

Design, G.B.S.; Conceptualization, G.B.S. and M.Č. and J.S.N.; Methodology, M.Č. S.J.N and H.S.; Samples provision, M.V., J.H., K.S., J.L. R.A.R.; Formal Analysis, M.Č and J.S.N.; Investigation, M.Č., J.S.N., N.D.; Data Curation, E.B.D. and J.S.N.; Writing, M.Č. and G.B.S.; Supervision, G.B.S.; Project Administration, G.B.S.; Funding Acquisition, G.B.S.

## Supplementary information

**Supplementary Figure 1 – Differential expression of ncRNAs in aging and AD**

Volcano plot of DE miRNAs with *P* value cut-off drawn at equivalent of FDR adjusted *P*=0.05 and fold-change cut-off drawn at ±1.5, heatmap of individual ncRNAs expression and PCA of miRNAs expression based on experimental groups are shown for all types of ncRNA, including **(a)** snoRNAs, **(b)** Box C/D fragments, **(c)** Box HACA fragments.

**Supplementary Figure 2 – The medianRank and Zsummary statistics of the module preservation**

**(a)** The medianRank of the modules close to zero indicates a high degree of module preservation. **(b)** The preservation Zsummary with dashed blue and green lines indicating the thresholds Z = 2 (low to moderate evidence of conservation) and Z = 10 (strong evidence of conservation), respectively.

**Supplementary Figure 3 – miRNA-target interactions of AD-related purple module miRNAs**

Specific miRNA-target interaction network of purple module showing miRNAs related to AD and their key protein targets (stars label miRNAs targeting APP).

**Supplementary Figure 4 – Characterization of miRNAs in AD and other neurodegenerative diseases**

Volcano plots show DE and WGCNA module membership of miRNAs linked to AD, PD, ALS, and FTD by select previous studies (*P* value cut-off drawn at equivalent of FDR adjusted *P*=0.05 and fold-change cut-off drawn at ±1.5).

**Supplementary Figure 5 – MiRNA relationships in aging and AD**

Schematic representation of differences in miRNA expression during aging in individual subclusters of miRNAs DE in all pairs of experimental groups (N=89). Dashed vertical lines connect pairs of schematics showing inverse versions of expression ratios within healthy old individual and patients with AD (relative to up-and down-regulation). Table shows individual subclusters, their change in expression within the studied groups and number of miRNAs belonging to each subcluster.

