## Supplementary figures and images for "Missorting of Plasma miRNAs in Aging and Alzheimer’s Disease"

### Fig. S1

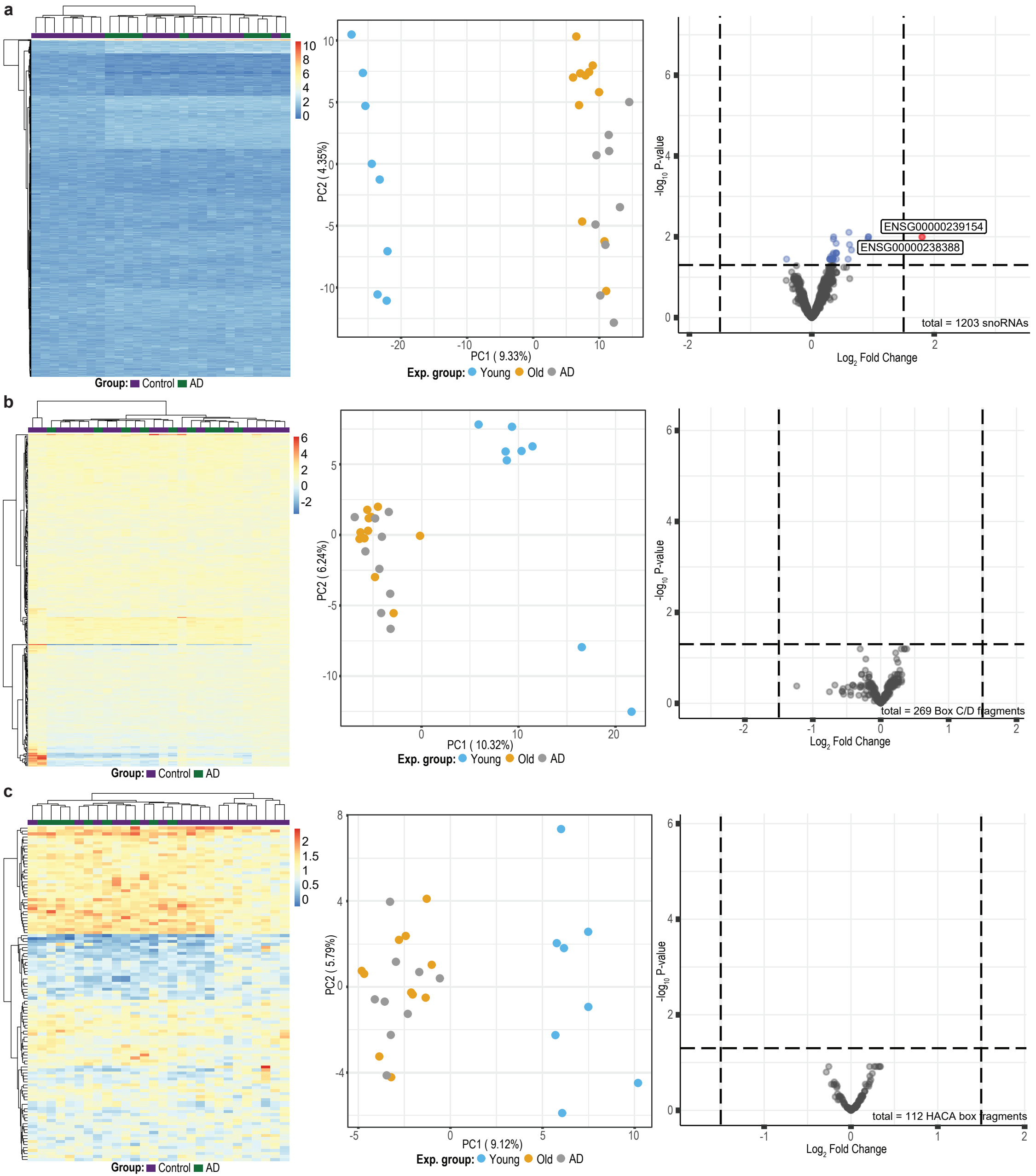

### Fig. S2

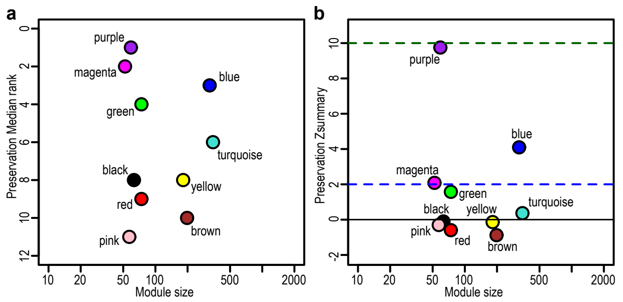

### Fig. S3

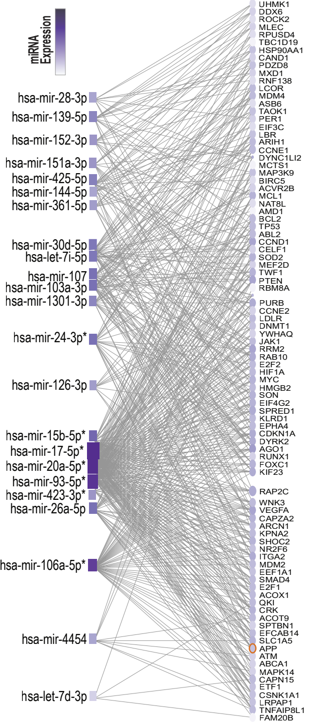

### Fig. S4

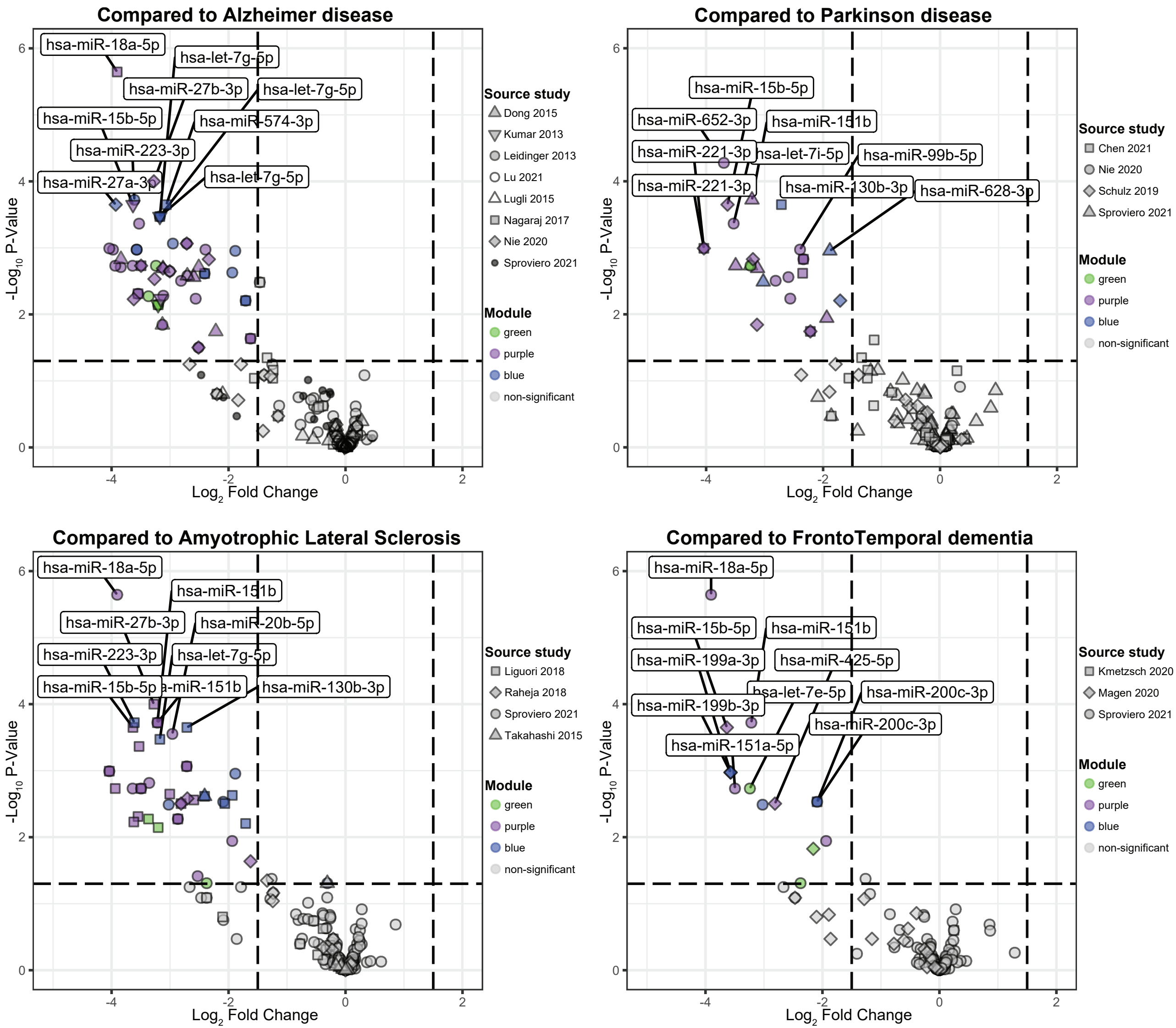
