## Supplementary material for "Missorting of Plasma miRNAs in Aging and Alzheimer’s Disease": Fig. S5

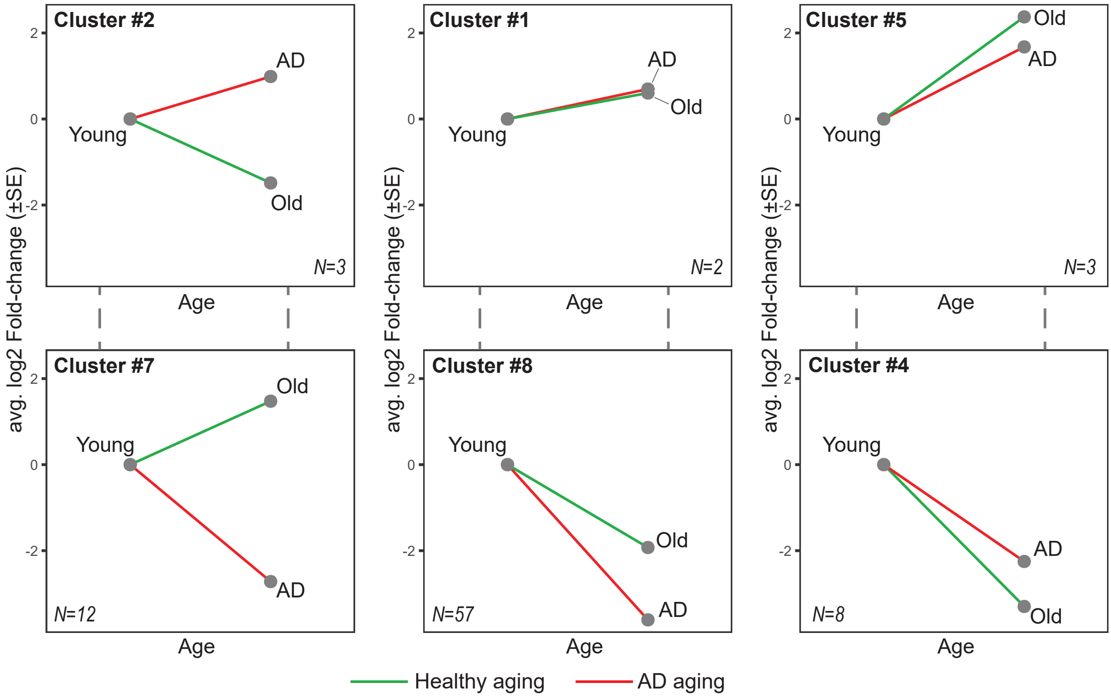


| Subcluster | Old ct Young | AD ct Young | AD ct Old | # miRNAs |
|---|---|---|---|---|
| Cluster #1 | up | up | up | 2 |
| Cluster #2 | down | up | up | 3 |
| Cluster #4 | down | down | up | 8 |
| Cluster #5 | up | up | down | 3 |
| Cluster #6 | down | up | down | 3 |
| Cluster #7 | up | down | down | 12 |
| Cluster #8 | down | down | down | 57 |
